# Safety of the fourth COVID-19 BNT162b2 mRNA (second booster) vaccine

**DOI:** 10.1101/2022.06.07.22276117

**Authors:** Matan Yechezkel, Merav Mofaz, Tal Patalon, Sivan Gazit, Erez Shmueli, Dan Yamin

## Abstract

COVID-19 remains a global concern due to vaccine protection waning and the emergence of immune-evasive variants. While the effectiveness of a second booster vaccine dose (i.e., fourth inoculation) is well proven, its safety has yet to be fully understood, and vaccine compliance remains low. We conducted a prospective observational study to compare the short-term effects of the first and second BNT162b2 mRNA COVID-19 vaccine booster doses. 2,019 participants received smartwatches and filled in a daily questionnaire on systemic reactions to the vaccine. We found substantial changes from baseline levels in the 72 hours post-vaccination with the second booster in both self-reported and physiological reactions measured by the smartwatches. However, no significant difference in reactions was observed between the first and second boosters. We also found that participants who experienced more severe reactions to the first booster tended to likewise experience more severe reactions to the second booster. Our work supports the safety of the second booster from both subjective (self-reported questionnaires) and objective (physiological measurements) perspectives.

## Introduction

Since its emergence in December 2019, SARS-CoV-2 (COVID-19) remains a global threat. Of particular concern is the apparent constant improvement in the ability of the virus to reinfect by evading the host immune system, despite previous infections with the virus and vaccinations. Specifically, the recent COVID-19 variant of concern (VOC), Omicron (B.1.1.529), features a substantial number of mutations in the receptor-binding domain of the spike protein, which enhances its replication and transmissibility ^1,2^. Indeed, there is growing evidence of reinfections with Omicron subvariants, including BA.2, BA2.12.1, BA.4, and BA.5, even among individuals previously infected with the first Omicron variant ^3–5^. Likewise, a rapid waning of COVID-19 vaccine protection has been observed as new VOCs appear ^6,7^. For example, recent evidence suggests that the BNT162b2 mRNA vaccine’s effectivity in decreasing morbidity declines from 67.2-73.9% to 45.7-64.4% within three months from booster dose administration ^8^. (For the sake of clarity, the BNT162b2 vaccine is administered in two doses, three weeks apart. The first booster dose is, in fact, a third dose of the vaccine, and the second booster dose, a fourth dose of the vaccine.)

Despite the demonstrated utility of the COVID-19 vaccines, there has been notable public reluctance to get vaccinated. Recent studies demonstrate that COVID-19 vaccine hesitancy is mainly motivated by safety concerns rather than efficacy considerations ^9,10^. A case in point is the second booster vaccine. The rapid spread of the Omricon variant and the sharp rise in Omicron-associated hospitalizations led the Israeli government to initiate, on December 30, 2021, a world-leading second mRNA booster vaccine campaign. The campaign focused on individuals at high risk and was expanded three days later to all individuals 60 years of age and older ^11^. Several countries have since followed suit. The US Centers for Disease Control and Prevention (CDC), for example, issued on March 29, 2022, a recommendation that individuals over 50 years of age and immunocompromised patients should receive a second mRNA booster dose ^12^. Yet, only 21.5% of eligible individuals had followed the CDC’s recommendation by May 27, 2022 ^13^. This reluctance may be, at least in part, because, while the second BNT162b2 booster dose was shown to be effective in preventing the severe outcomes of COVID-19 ^14,15^, its safety is yet to be fully understood. This knowledge gap stems from the lack of sufficient continuous and detailed monitoring and surveillance of vaccinated individuals, a gap that has thus far been filled by non-scientific, rather speculative theories ^16^. The need to rectify this situation is highlighted by the CDC’s decision to award the Global Vaccine Data Network US$5.5 million to investigate vaccine safety data in a prospective manner ^17^.

Several recent studies have demonstrated the excellent performance of wearable sensors such as smartwatches in detecting physiological changes following vaccination, as they can continuously monitor reactions in an objective manner ^18–21^. Impressively, they have been shown to be even more sensitive than humans in detecting vaccine reactions ^18,19^, even in individuals who did not report any reactions following vaccination ^18^. It has been shown, for example, that changes in heart rate measures following vaccination correlate with the severity of vaccine reactions and could accurately determine when participants’ heart rates returned to baseline levels ^21^.

Here, we compare the short-term effects of the first and second BNT162b2 mRNA COVID-19 vaccine booster doses. Specifically, we followed 2,019 participants as part of the PerMed observational prospective study ^18–20^. Our study participants received Garmin Vivosmart 4 smart fitness trackers and filled in a daily questionnaire on systemic reactions via a dedicated mobile application, from at least seven days before vaccination and 30 days thereafter. The smartwatch continuously monitored several physiological measures, including heart rate levels. In addition, participants gave consent to access their EMRs, which included information regarding the exact hour of vaccine administration. This rich and versatile data allowed us to examine the safety of the second booster vaccine, gleaned from participants’ objective physiological measures and medical records together with their subjective reports.

## Results

Of the 2,019 participants who received either the first or second booster, 1,024 (50.72%) were women, and 994 (49.23%) were men (one participant did not specify his/her gender). Their age ranged between 19 and 89 years, with a median age of 52 years (Table 1). In the study cohort, 1,560 (77.27%) had a body mass index (BMI) below 30, and 462 (22.88%) had at least one specific underlying medical condition (Table 1). Around 30% of the participants, 615, received a second booster dose during the study period. Out of them, 385 also received the first booster dose during the study period. The distributions of gender were relatively invariable across the recipients of the first and second booster doses. In contrast, the distributions of age and underlying medical condition differed, due to the eligibility criteria for the second booster dose (Table 1).

**Table 1.** Characteristics of the participants

| Characteristic | First Booster<br>(N=1,789) | Second Booster<br>(N=615) | First and Second<br>Booster<br>(N=385) |
| --- | --- | --- | --- |
| **Gender** |  |  |  |
| Male | 48.69% (918) | 49.92% (307) | 47.79% (184) |
| Female | 51.31% (871) | 49.92% (307) | 52.21% (201) |
| Unspecified | 0.00% (0) | 0.16% (1) | 0.00% (0) |
| **Age group** |  |  |  |
| 18-29 yr | 15.43% (276) | 3.90% (24) | 2.60% (10) |
| 30-39 yr | 19.90% (356) | 7.80% (48) | 5.19% (20) |
| 40-49 yr | 11.29% (202) | 7.80% (48) | 3.38% (13) |
| 50-59 yr | 24.04% (430) | 16.10% (99) | 16.62% (64) |
| 60-69 yr | 20.18% (361) | 41.30% (254) | 43.12% (166) |
| ≥ 70 yr | 9.17% (164) | 23.09% (142) | 29.09% (112) |
| **Body-mass index\*** |  |  |  |
| < 30.0 | 77.19% (1381) | 73.17% (450) | 70.40% (271) |
| ≥ 30.0 | 20.51% (367) | 23.74% (146) | 24.93% (96) |
| Unspecified | 2.30% (41) | 3.09% (19) | 4.67% (18) |
| **Comorbidities\*\*** |  |  |  |
| Yes | 21.86% (391) | 39.35% (242) | 44.42% (171) |
| No | 75.63% (1,351) | 56.42% (347) | 50.13% (193) |
| Unspecified | 2.51% (45) | 4.23% (26) | 5.45% (21) |

\* The body-mass index is the weight in kilograms divided by the square of the height in meters
\*\* Comorbidities are defined as having at least one of the following: hypertension, diabetes, heart disease, chronic lung disease, immune suppression, cancer, and renal failure.

### Short-term effects of the second booster

For 508 of the 615 participants who received the second booster, we had sufficient smartwatch data to compare their objective physiological indicators’ levels (i.e., heart rate and heart rate variability (HRV)) from seven days prior to the administration of the second booster dose (the baseline period) with those in the seven days after the vaccination. We identified a considerable rise in both the heart rate (Figure 1A) and HRV-based stress (Figure 1B) indicators in the first 48 hours following the administration of the second booster compared to baseline levels (p-value < 0.001). These changes faded after the initial 72 hours, with measurements returning to their baseline levels.

**Fig. 1.**
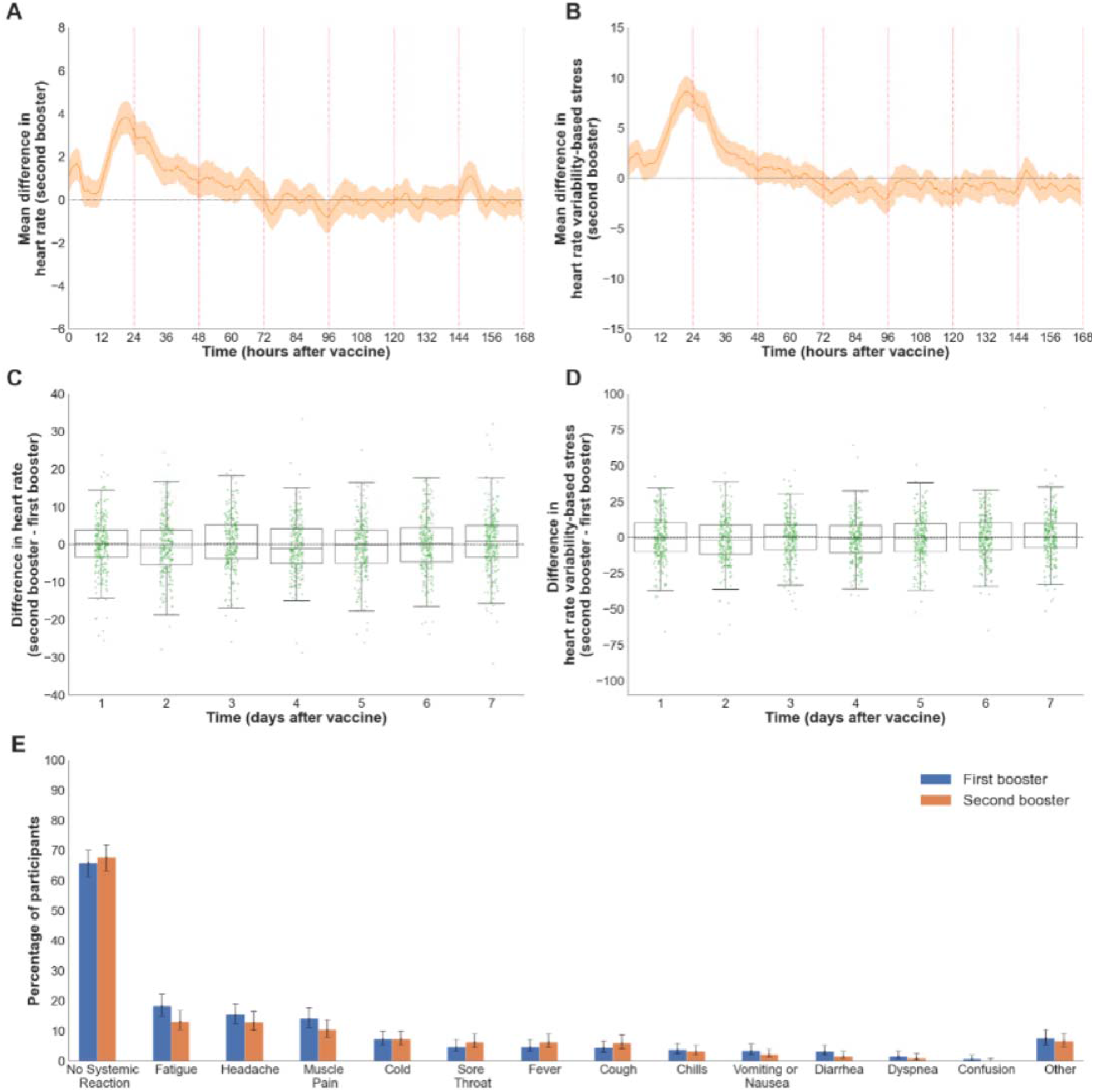
Self-reported and physiological reactions to the second booster dose compared to the first booster dose. (**A-B**) Reactions to the second booster as recorded by the smartwatches. The figures show the mean difference between the baseline and post-vaccination period in (A) heart rate (n=507) and (B) heart rate variability-based stress (n=505). Mean values are depicted as solid lines, and 90% confidence intervals are presented as shaded regions. (**C-D**) A comparison of the reactions recorded by the smartwatches between the first and second boosters. The figures show boxplots of the differences between the daily mean changes in smartwatch indicators (each change is between the post vaccination and baseline periods) of the second and first boosters (C) heart rate (n=301) and (D) heart rate variability-based stress (n=297). Each green dot represents a single participant. (**E**) A comparison of the reactions reported by participants between the first and second boosters (n=316). The bars represent the percentage of participants who reported a given reaction. Error bars represent 90% confidence intervals.

Similarly, for 553 of the 615 participants who received the second booster, we had sufficient questionnaire data to assess the extent of reactions reported following the second booster vaccine dose (Figure S1). Specifically, 65.5% (90% CI: 62.1-68.7%) of the participants did not report any new symptom after receiving the second booster dose. The most frequent reported reactions were fatigue, headache, muscle pain, cold, and a sore throat. These reactions faded in nearly all participants within three days.

### Pairwise comparative analysis of the first and second boosters

We conducted a pairwise comparative analysis between reactions to the two boosters for participants who received the two boosters during the study period (Figure 1C-E). For the two physiological indicators, heart rate and HRV-based stress, we first calculated for each participant the daily mean change between the post-vaccination and baseline periods, separately for the first and second boosters. Then, we calculated the difference between these two means for each participant and each of the seven days after inoculation (Figure 1C, D). Our analysis revealed no significant difference in both physiological measures between the second and first booster (one-sample t-test, p-value=0.099). Similarly, an analysis of the self-reported reactions post-vaccination revealed that the extent of systematic reactions reported following the second booster dose was similar to those observed following the first booster dose (McNemar’s statistic=2.753, p-value=0.097) (Figure 1E). For example, out of 316 participants (with sufficient questionnaire data of the first and second boosters), 67.7% (90% CI: 63.2-71.9%) did not report any new symptom after receiving the second booster dose, compared to 65.8% (90% CI: 61.3-70.0%) after the first booster dose. Moreover, the most frequently reported reactions (i.e., fatigue, headache, muscle pain, fever, and cold) were similar after the first and second booster doses.

We also explored the association between the severity of the physiological reactions to the first booster vs. the second booster. We fitted a multiple linear regression model to explore the effect of the magnitude of the physiological reaction to the first booster on the magnitude of the physiological reaction to the second booster, controlling for other explanatory variables (age, and time from first to second booster). Interestingly, we found no association between the magnitudes of reactions in heart rate and HRV-based stress (p-value=0.109, p-value=0.358, respectively) (Table S1).

We also categorized each participant’s self-reported reactions after each booster administration into three categories, “Severe Reaction”, “Mild Reaction”, and “No Reaction”, in line with the CDC guidelines (see Methods). To isolate the effect of the severity of reactions following the first booster from other possible explanatory variables (age, and time from first to second booster), we fitted an ordinal logistics regression model (Table S2). We found that the association between the severity of reaction following the first and second boosters was significant (p-value<0.001).

## Discussion

Our comparison of data on individuals who received the first vs. second booster of the BNT162b2 mRNA COVID-19 vaccine dose revealed no significant difference in the peak and time until return to the baseline of the physiological indicators and self-reported reactions. Our findings further suggested that reactions reported following the second booster dose administration were significant in terms of both the self-reported adverse events and the physiological reactions observed by the smartwatches. These findings were consistent for participants for whom we had information on both doses (i.e., in the paired-wised analysis).

Importantly, our analysis of the self-reported reactions found a significant linear association between the severity of the symptoms of the first and the second booster, while the objective data (i.e., physiological indicators) indicated an insignificant linear association. This could be attributed to variations between participants in terms of their own sensitivity to changes in their body. This observation is consistent with a previous study that indicated that self-reported reactions were more apparent for women than for men. However, the objective reactions, detected by the smartwatches did not differ between the genders ^18^. Regardless of the actual reason, this discrepancy highlights the importance of also accounting also for objective measures to determine safety.

While our cohort size is relatively reasonable compared to other studies using wearable devices, it has a rather large paired-wise sample ^18,19,21^. In particular, most of the participants in our second booster analysis are over 50 years old, in line with current Israeli and CDC vaccine administration guidelines ^22,23^. This age group is characterized by a higher rate of underlying medical conditions and might be more sensitive to changes caused by vaccine reactions. Thus, if guidelines are changed to include all individuals aged 18 years and above, our findings may not be generalized.

As a final remark, we stress that while our study supports the safety of the fourth dose, there remains a stark and growing imbalance in COVID-19 vaccine coverage worldwide. Nearly all middle- and low-income countries have not secured the first booster dose for their citizens. Although mutations occur by chance, multiple compounding factors of disadvantage may facilitate the spread of variants in low-resource settings ^24^. Thus, although it seems in the interest of high-income countries to prioritize vaccinating their own population with a second booster, it is a moral obligation and in their personal self-interest to support equitable vaccine distribution.

In conclusion, our analyses show no significant differences between the second and first booster doses in terms of the peak and time until the return to baseline of the physiological indicators and the self-reported reactions. Additionally, the severity of systematic reactions after the second booster is associated with the severity of reactions following the first booster dose. Thus, our findings further highlight the short-term safety of the BNT162b2 second booster dose, as reflected in both subjective and objective data.

## Data Availability

Researchers interested in obtaining an aggregated version of the data sufficient to reproduce the results reported in this paper should contact the corresponding author.

## Acknowledgments

This research was supported by the European Research Council (ERC) project #949850 and the Israel Science Foundation (ISF), grant No. 3409/19, within the Israel Precision Medicine Partnership program.

## Authors’ contributions

Conception and design: DY, MY, ES. Collection and assembly of data: DY, TP, SG, ES. Analysis and interpretation of the data: MY, MM, DY, ES. Statistical expertise: MY, MM, DY, ES. Drafting the article: MY, DY, ES. Critical revision of the article for important intellectual content: DY, ES. Final approval of the article: All authors. Obtaining funding: DY, ES.

## Code and Data availability

Researchers interested in obtaining an aggregated version of the data sufficient to reproduce the results reported in this paper should contact the corresponding author. Statistical code will be available after acceptance.

## Methods

### Ethical approval

Before participating in the study, all the subjects were advised, both orally and in writing, as to the nature of the study and gave written informed consent. The study was approved by Maccabi Health Service’s Helsinki institutional review board, protocol number 0122-20-MHS.

### Study design and participants

The PerMed prospective observational study includes 4,698 participants (18+ years of age) who were recruited between November 1, 2020, and March 30, 2022 (see also detailed information in previous studies –^18,20^). From this cohort, the 2,019 participants who received either the first or second BNT162b2 mRNA COVID-19 vaccine booster dose after joining the study served as the base dataset for our analysis. Of these 2,019 participants, during the study, 1,789 received a first booster dose, 615 a second booster dose, and 385 received both the first and second booster doses. Notably, all participants in our study received the BNT162b2 mRNA vaccine.

We employed a professional survey company to recruit participants and ensure they adhere to the study requirements. Participant recruitment was performed via advertisements on social media and word-of-mouth. Each participant signed an informed consent form after receiving a comprehensive explanation on the study. Then, participants completed a one-time enrollment questionnaire, were equipped with Garmin Vivosmart 4 smartwatches, and installed two applications on their mobile phones: (1) the PerMed application ^18–20^, which collects daily self-reported questionnaires, and (2) an application that passively records smartwatch data. Participants were asked to wear their smartwatches as much as possible. The survey company ensured that participants’ questionnaires were filled at least twice a week, that their smartwatches were charged and properly worn, and that any technical problems with the mobile applications or smartwatch were resolved. Participants were monitored through the mobile application and smartwatches for a period of at least 37 days, starting seven days before vaccination. Participants also granted full access to their EMR data.

We implemented several preventive measures to minimize participant attrition and discomfort as a means to improve the quality, continuity and reliability of the collected data. First, each day, participants who did not fill their daily questionnaire by 7 pm received a reminder notification through the PerMed application. Second, we developed a dedicated dashboard that allowed the survey company to identify participants who repeatedly neglected to complete the daily questionnaire or did not wear their smartwatch for extended periods of time; these participants were contacted by the survey company (either by text message or phone call) and encouraged to better adhere to the study protocol. Third, to strengthen participants’ engagement, a weekly personalized summary report was generated for each participant, which was available inside the PerMed application. Similarly, a monthly newsletter with recent findings from the study and useful tips regarding the smartwatch’s capabilities was sent to the participants.

### Self-reported questionnaires

Upon enrollment in the study, we collected information on participants’ gender, age and underlying medical conditions. The list of underlying medical conditions consisted of hypertension, diabetes, heart disease, chronic lung disease, immune suppression, cancer, renal failure and BMI>30 (BMI is computed as the weight in kilograms divided by the square of the height in meters). After the enrollment, participants used the PerMed mobile application ^18–20^ to fill out a daily questionnaire. The questionnaire allowed participants to report on potential clinical symptoms, as observed in the BNT162b2 mRNA Covid-19 clinical trials ^25^, with an option to add other symptoms as free text.

### The smartwatch

Participants were equipped with Garmin Vivosmart 4 smart fitness trackers. Among other features, the smartwatch provides all-day heart rate (HR) and heart rate variability (HRV)-based stress ^26^. The smartwatch’s optical wrist HR monitor is designed to continuously monitor a user’s heart rate. The frequency at which heart rate is measured varies and may depend on the level of activity of the user: when the user starts an activity, the optical HR monitor’s measurement frequency increases.

Since HRV is not easily accessible through Garmin’s application programming interface (API), we use Garmin’s stress level instead, which is calculated based on HRV. Specifically, the device uses heart rate data to determine the interval between each heartbeat. The variable length of time between each heartbeat is regulated by the body’s autonomic nervous system. Less variability between beats correlates with higher stress levels, whereas an increase in variability indicates less stress ^27^. A similar relationship between HRV and stress was also seen in ^28,29^. Examining the data collected in our study, we identified an HR sample roughly every 15 seconds and an HRV sample every 180 seconds.

Of note, while the Garmin smartwatch provides state-of-the-art wrist monitoring, it is not a medical-grade device, and some readings may be inaccurate under certain circumstances, depending on factors such as the fit of the device and the type and intensity of the activity undertaken by a participant ^30–32^.

### Data preprocessing and inclusion criteria

The questionnaire data were preprocessed by manually categorizing any self-reported symptom entered as free text. If participants filled out the questionnaire more than once in one day, the last entry from that day was used in the analysis. Smartwatch data were preprocessed as follows. First, we computed the mean value of each hour of data. We then performed a linear interpolation to impute missing hourly means. Lastly, we smoothed the data by calculating the five-hour moving average.

For each participant and each of the two booster doses, we defined the 7-day period prior to vaccination as the baseline period. For the analyses involving self-reported questionnaires, we included participants who submitted at least one questionnaire during the baseline period and at least one questionnaire during the seven days post vaccination. The two questionnaires were required for understanding the appearance of new reactions following vaccination. For the analyses involving smartwatch indicators, we included participants who had at least one overlapping period of data (i.e., same day of the week and same hour during the day) during their baseline and post vaccination periods. The overlapping periods were required for computing the change in indicator values between the baseline and post-vaccination periods.

### Statistical analysis

To compare the changes in smartwatch indicators over the seven days (168 hours) post vaccination, with those of the baseline period, we performed the following steps. First, for each participant and each hour during the seven days post-vaccination, we calculated the difference between that hour’s indicator value and that of the corresponding hour in the baseline period (keeping the same day of the week and same hour during the day). Then, we aggregated each hour’s differences over all participants to calculate a mean difference and the associated 90% confidence interval, which is analogous to a one-sided t-test with a significance level of 0.05. To determine the statistical significance of daily differences between the baseline and post vaccination period, we calculated the mean daily difference for each participant and then used a one-sample t-test for each day.

To understand the extent of new reactions post vaccination, we first noted any pre-existing clinical symptoms reported in the last completed questionnaire during the baseline period. Next, we calculated the percentage of participants who reported new (i.e., not pre-existing) systemic reactions in the 7-day period after vaccination. For each reaction, we used a beta distribution to determine a 90% confidence interval..

#### Pairwise comparative analysis

In this set of analyses, we considered only participants who received both the first and second boosters during the study period.

To determine the statistical significance of the difference between the physiological reactionsto the second booster vs. the first booster, we first calculated for each participant the daily mean change between the post vaccination and baseline periods separately for the first and second boosters. Then, we calculated the difference between these two means for each participant. Finally, we used a one-sample t-test to assess whether the mean difference of daily changes was significantly different from zero. To determine the statistical significance of difference between the extent of reported reactions to the second booster vs. the first booster, we performed a McNemar sign test.

To examine the relations between the intensity of physiological reactions to the second booster vs. the first booster, controlling for other explanatory variables (age, and interval between boosters), we fitted two multiple linear regressions ^33^, one for each of the smartwatch indicators. We defined the intensity of the reaction as the mean change in the indicator’s values in the first 48 hours post vaccination compared to baseline values:

To perform a similar analysis for the self-reported reactions, we first had to define their severity. For participants who reported feeling hot and who recorded their temperature, we classified the temperature as above 37.5ºC (fever) or below 37.5ºC (feeling hot); if the participant did not record their temperature, we classified the temperature as below 37.5ºC. In line with the CDC and the Pfizer clinical trial^25,34^, we categorized symptoms as follows:

- Mild symptoms: abdominal pain, feeling hot, back or neck pain, feeling cold, muscle pain, weakness, headache, dizziness, vomiting, sore throat, diarrhea, cough, leg pain, ear pain, loss of taste and smell, swelling of the lymph nodes, fast heartbeat, and hypertension;
- Severe symptoms: chest pain, dyspnea (shortness of breath), fever, confusion, and chills.

Consequently, participants were either classified as having “No Reaction,” a “Mild Reaction,” or a “Severe Reaction”, based on their most severe symptom reported in the seven days after each vaccination. Thus, if a participant reported one severe symptom for one day and mild symptoms for all three days after vaccination, the participant was classified as having a severe reaction. Participants could be categorized into different severity groups after each booster dose.

Then, to examine the relation between the severity of reported reactions to the second booster vs. the first booster, controlling for other explanatory variables (age, and interval between boosters), we fitted an ordinal logistic regression ^35^.

## Supplementary Materials for

## Additional Results

### Self-reported reaction to the second booster dose

The majority of participant did not report any new systematic reaction during the 7-day period postvaccination. The most frequent reported reactions were fatigue, headache, muscle pain, cold, and a sore throat. These reactions faded in nearly all participants within three days (Figure S1).

**Figure S1.**
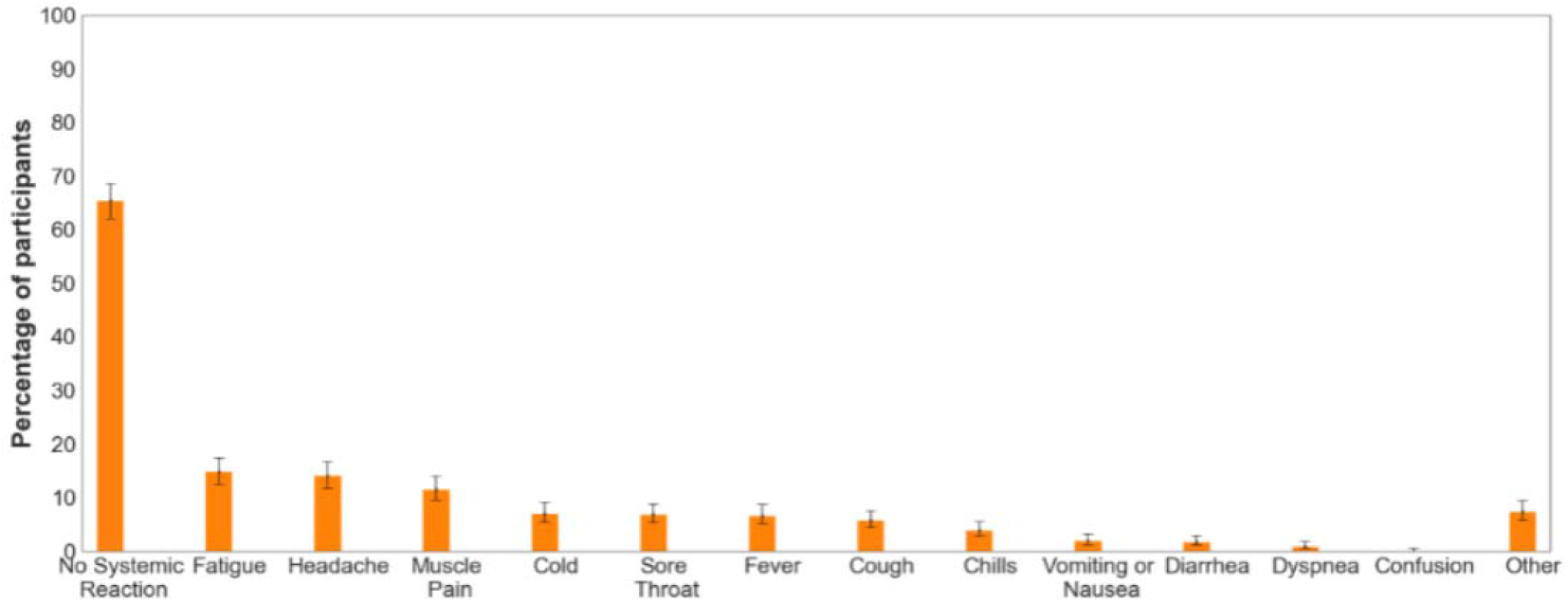
Self-reported reaction to the second booster dose. The bars represent the percentage of participants who reported a given reaction. Error bars represent 90% confidence intervals (n=553).

### Multiple linear regression model for physiological indicators

Our multiple linear regression model revealed an insignificant linear association between the intensity of physiological reactions 48 hours following the second booster and the explanatory variables: age, the time interval between boosters, and the intensity of physiological reactions 48 hours following the first booster (Table S1).

**Table S1.** Multiple linear regression model results for heart rate and HRV-based stress

| **Variable** | **Value (95% CI)** | **P-value** |
| --- | --- | --- |
| **<u>Hear rate linear regression</u>** |  |  |
| F-statistic | 1.005 | 0.391 |
| **Coefficients** |  |  |
| Intercept | 0.406 (0.296-0.515) | <0.001 |
| Time between boosters | 0.006 (-0.106-0.119) | 0.910 |
| Age | 0.038 (-0.062-0.138) | 0.450 |
| Heart rate after first booster | 0.120 (-0.027-0.267) | 0.109 |
| **<u>HRV-based stress linear regression</u>** |  |  |
| F-statistic | 1.447 | 0.230 |
| **Coefficients** |  |  |
| Intercept | 0.664 (0.580-0.747) | <0.001 |
| Time between boosters | 0.023 (-0.072-0.117) | 0.635 |
| Age | -0.067 (-0.151-0.016) | 0.113 |
| HRV-based stress after first booster | 0.045 (-0.52-0.143) | 0.358 |

### Ordinal logistics regression model for self-reported reactions

Our ordinal logistics regression model revealed a significant association between the severity of reported reactions to the second booster and the explanatory variables: age, the time interval between boosters, and the severity of reported reactions to the first booster (Table S2).

**Table S2.** Ordinal logistics regression model results for the severity of self-reported reactions

| **Coefficient** | **Value (95% CI)** | **P-value** |
| --- | --- | --- |
| Time between boosters | -0.022 (-0.038-(-0.006)) | 0.006 |
| Age | -0.044 (-0.065-(-0.024)) | <0.001 |
| Severity of reported reactions to the first booster | 1.322 (0.939-1.706) | <0.001 |
| Intercept: Health/Mild | -3.722 (-6.738-(-0.706)) | 0.016 |
| Intercept: Mild /Severe | 0.837 (0.629-1.046) | <0.001 |

